# Impact of disease-modifying therapies in adults with concomitant psoriatic and metabolic liver disease with integrated immunoprofiling

**DOI:** 10.64898/2026.07.06.26357384

**Authors:** Shannon Gunawardana, Lija James, Charlie Diamond, Anneli Andersson, Alessandro Fichera, Jiaqi Li, Sinibaldo Romero Arocha, Moustafa Attar, Hussein Al-Mossawi, Paul Klenerman, Helena Thomaides-Brears, Alexander J Clarke, Laura C Coates

## Abstract

Psoriatic disease (PsD) is associated with metabolic dysfunction-associated steatotic liver disease (MASLD), but the hepatic effects of biologic therapies are unclear. We evaluated paired liver MRI and multi-modal immunoprofiling in PsD patients initiating new systemic therapy.

COLIPSO is a prospective cohort of adults with moderate-to-severe psoriasis or psoriatic arthritis (PsA) starting a new conventional synthetic or biologic disease-modifying antirheumatic drug (DMARD). Liver MRI was performed at baseline and ∼6 months. A subset of participants with PsA underwent peripheral blood flow cytometry and single-cell RNA sequencing (scRNAseq). Primary outcomes were within-subject change in quantitative MRI measures of liver disease activity and fat content (iron-corrected T1 [cT1] and proton density fat fraction [PDFF]). Bayesian models were used.

Thirty-five participants (mean age 50±13 years; 61% male) were followed for ∼29 weeks. Baseline disease activity was moderate (mean DAPSA 29) and 40% had MASLD. IL-17 inhibitors (IL-17i) improved PDFF (−1.58 ± 1.61%) and cT1(−43.6 ± 52.7ms), whereas TNFi showed little change. Compared with csDMARD, IL-17i improved PDFF (probability of direction [pd] 89%) and cT1 (pd 93%), which was not seen with TNFi. Flow cytometry (n=17) linked baseline γδ and ThGM-CSF T-cell abundance with cT1 and PDFF. scRNAseq highlighted baseline transcriptomic signatures in MAIT cells associated with cT1 and PDFF. Naïve T-cell RNA signatures at baseline were associated with MRI improvements.

In PsD, only IL-17i were associated with improved liver disease in addition to improving clinical PsD outcomes. T-cell subtypes bridging innate and adaptive immunity were associated with liver disease features.

**Key messages:**

- IL-17 inhibition was associated with a significant reduction in hepatic steatosis (PDFF) and liver inflammatory disease activity (cT1).
- Baseline γδ and ThGM-CSF T cell abundance was associated with cT1 and PDFF, and γδ T cells and cT1 and PDFF co-varied over follow up. Distinct gene expression signatures in MAIT and naïve CD4 T cells at baseline were associated with liver MRI parameters.
- Our results support larger mechanistic and clinical studies to test if IL-17 pathway inhibition improves liver disease in PsD, and whether treatment decision algorithms should account for metabolic liver disease.

## Introduction

Psoriatic disease (PsD) is a chronic inflammatory condition with systemic implications that extend beyond skin and joint involvement. Psoriatic arthritis (PsA) is clinically heterogeneous and may affect the peripheral and axial joints, entheses, digits (dactylitis), and nails. Psoriasis (PsO) prevalence in Western Europe ranges from 1-5%, with 20-30% of these individuals developing PsA. [1, 2]

There is a well-established link between PsD and metabolic dysfunction-associated steatotic liver disease (MASLD).[3–5] MASLD is defined by cardiometabolic risk and excess liver fat, and can progress to metabolic dysfunction-associated steatohepatitis (MASH), which is additionally characterised by disease activity (steatosis, ballooning, and inflammation) and can lead to further structural changes and organ injury.[6] Without intervention, MASH can progress to advanced fibrosis, cirrhosis, and ultimately end-stage liver disease or hepatocellular carcinoma; notably these individuals carry a significantly elevated risk of major adverse cardiovascular and liver-related events.[7, 8]

Multiparametric magnetic resonance imaging (mpMRI) techniques of the abdomen, including measurement of liver iron-corrected T1 (cT1) and proton density fat fraction (PDFF), are increasingly utilised in clinical practice, as supported by real-world evidence.[9] These techniques provide a comprehensive, non-invasive assessment of both liver disease activity and fat content. Unlike biopsy, mpMRI captures the heterogeneity of liver pathology across the entire organ, eliminates sampling error, and allows for safe, repeatable monitoring over time, making it a validated and reliable non-invasive alternative to biopsy in clinical practice.[10] PDFF correlates with histologically graded steatosis across the spectrum of MASH.[11–13] cT1 signal measures regional tissue water content and tissue composition, reflecting alterations in liver tissue water, increased steatosis, and extracellular matrix microenvironment associated with fibrogenic, parenchymal remodelling, and inflammatory processes. cT1 correlates with ballooning, steatosis, fibrosis, and the NAFLD activity score (NAS) from histology.[14–16]

Both PsD and MASLD are characterised by chronic systemic inflammation. PsO and PsA are T cell mediated diseases associated with proinflammatory cytokines including tumour necrosis factor (TNF), interleukin (IL)-17 and IL-23.[17, 18] Intriguingly IL-17-signalling is upregulated is MASLD, and is thought to contribute to inflammation and progression of liver disease, through synergistic effects with other cytokines such as TNF and IL-1β.[19, 20] The identification of these shared inflammatory cytokines and pathways suggests the potential for therapies that offer benefits across both PsD and MASLD. To date, investigations into MASLD and liver toxicity in PsD have focused almost exclusively on methotrexate, with limited data addressing the effects of biologic therapies, such as, TNF and IL-17-inhibitors.[21–23] There is a need for prospective data to understand the interplay between psoriatic disease activity, biologic treatments, immunopathogenesis and liver health in psoriatic disease.

Therefore, in this exploratory study we used a multimodal approach to assess changes in mpMRI liver parameters (cT1, indicating liver disease activity and PDFF, indicating liver fat), and their association with T cell immunophenotype and clinical metrics in adults with PsD, switching disease-modifying treatment. We evaluated the influence of biologic and non-biologic therapies on MASLD progression and identified immunological markers of treatment response in clinical and liver outcomes.

## Methods

### Study design

The Co-Prevalence of Liver Disease in Psoriatic Disease (COLIPSO) study (London - Bromley Research Ethics Committee, REC: 20/LO/0616, IRAS ID: 274846) was a real-world, multi-centre study adopting a prospective longitudinal cohort study design in individuals with PsD, as reported previously.[24] This study reports on participants who attended both a baseline and a follow-up visit 24-weeks later.

### Sampling and recruitment

Adults aged ≥18 years with PsD were enrolled from rheumatology and dermatology clinics at Oxford University Hospitals NHS Foundation Trust (Oxford, UK) and Leeds Teaching Hospitals NHS Trust (Leeds, UK) between 2021 and 2024. Inclusion criteria were a physician-confirmed diagnosis of moderate to severe psoriasis or psoriatic arthritis, which required initiation of a new systemic therapy, either a conventional synthetic DMARD (csDMARD) or a biologic DMARD (bDMARD). All enrolled subjects underwent thorough clinical assessment and MRI evaluation at the time of recruitment or within 7 days of starting a new treatment, and 6 months after starting new treatment. Exclusion criteria were contraindications to MRI; a diagnosis of autoimmune hepatitis, viral hepatitis, or Wilson’s disease; or any other medical condition deemed by the investigator to pose a safety risk or impair study participation.

### Data collection

At baseline and follow-up, the following data were collected: demographic and anthropometric measurements; medical and medication history, alcohol intake and disease duration, Psoriasis Area and Severity Index (PASI) score and percentage body surface area (BSA) affected by psoriasis, swollen joint count (SJC), tender joint count (TJC), prior and current use of DMARDs, patient reported health related quality of life outcomes, including pain and global visual analogue scales (VAS); liver function tests (bilirubin, albumin, alanine aminotransferase or alkaline phosphatase), Enhanced Liver Fibrosis (ELF) score, C-reactive protein (CRP) level and blood samples taken for additional laboratory analysis at baseline and follow up (flow cytometry, single-cell RNA sequencing, see Supplementary Methods). In addition, for adults with psoriatic arthritis (PsA), PsA Disease Activity Score (PASDAS) and Disease Activity in Psoriatic Arthritis (DAPSA) scores were recorded (based on 68 tender and 66 swollen joint counts, patient pain scores, global health and CRP), with DAPSA ≥ 28 defining severe PsA. Minimal Disease Activity (MDA) was used to define a state of remission.[25]

All participants underwent standardised, quantitative mpMRI (LiverMultiScan or CoverScan, Perspectum, Oxford) at both timepoints using clinical scanners: GE Signa Voyager 1.5 T (Chicago, IL, USA), Siemens Area 1.5 T (Forchheim, Germany) and Philips Achieva dStream 3 T (Amsterdam, Netherlands).[26] The image acquisition and processing were performed by trained MR technologists and radiographers blinded to clinical data, and all imaging datasets were centrally curated and quality controlled.

### Statistical analysis

Descriptive statistics (means, standard deviations, and sample sizes) were generated overall and by treatment group, defined by the new DMARD. Paired measurements were used to summarise within-subject changes from baseline to follow-up.

Normal values/reference ranges for MRI liver fat content and disease activity were defined as PDFF <5% and cT1 <800ms, respectively.[27, 28] MASLD was defined as fat accumulation in ≥5% of hepatocytes, in conjunction with at least one cardiometabolic risk factor, while explicitly excluding significant alcohol consumption or other secondary causes of liver fat accumulation. [5] MASH was defined by both elevation in liver fat content (≥5%) and disease activity (cT1 ≥ 800ms) and a cardiometabolic risk factor. These thresholds have previously been shown to be diagnostic of steatohepatitis in biopsy-paired datasets, with cT1 ≥ 875ms associated with risk of significant liver fibrosis.[27, 29, 30]

To quantify longitudinal changes and treatment-specific effects, for each outcome we fitted a Bayesian hierarchical (mixed-effects) Gaussian linear model with a patient-level random intercept to accommodate the within-patient correlation of the repeated measurements. The models included fixed effects for visit, medication group, and their interaction, along with covariates [age, sex, body mass index (BMI), and presence of psoriasis]. We used weakly informative priors, and all fixed effect coefficients were assigned independent normal distributions, and residual and between-patient standard deviations were given an exponential prior. The following model structure was used: outcome ∼ visit + treatment group + age + sex + BMI + (1 | patient).

For analysis of flow cytometry data at baseline, weakly informative priors were again specified for all parameters: normal priors centred at zero for regression coefficients, and half-normal priors for variance components. The intercept prior was centred on the observed outcome mean. For baseline data, the following model was used: standardized outcome ∼ standardized immune measure + age + sex + BMI + treatment group.

For the longitudinal mixed model, fixed-effect prior scales were computed from a base prior specification, then adaptively scaled by predictor spread and sample-size shrink factors with lower/upper clamps. Group-level (random-effect) prior scales were similarly adaptively shrunk and used as half-normal hyperpriors. This induces partial pooling across immune markers and regularization of weakly identified effects. The following model was used: standardized outcome ∼ 1 + visit + standardized immune measure (between patients) + standardized immune measure (within patients) + age + sex + BMI + group + visit × group + (0 + standardized immune measure (between patients) | standardized immune measure) + (1 | patient).

Posterior inference was performed using Markov chain Monte Carlo sampling implemented in the Julia package Turing, with four chains, 1000 warm-up iterations, and 2,000 posterior samples per chain (target acceptance probability = 0.95). Effects were summarised directly from the posterior by the posterior mean, the 95% credible interval (CrI; 2.5th–97.5th percentiles), and the probability of direction (pd), defined as the posterior probability that the effect takes its estimated sign.

## Results

Thirty-five individuals with PsD were recruited from secondary care, including 25 with PsA and 10 with PsO. These participants were stratified according to the DMARD class initiated at baseline: csDMARD (n = 15), TNFi (n = 15) and IL-17i (n = 5) (Supplementary Figure 1). At baseline, the mean age of the cohort was 49.5 years, 22/35 (63%) were male, and mean BMI was 28.9 kg/m^2^. Overall, 5/35 (14%) were lean, 18/35 (51%) were overweight and 12/35 (34%) had obesity (Table 1). Baseline inflammatory disease activity was moderate overall, with mean DAPSA 29, PASDAS 2.2, and PASI 4.4. Liver-related abnormalities were common: MASLD was present in 14/35 (40%) and MASH in 9/35 (25.7%). Mean baseline ELF was 8.7, ALT 34 IU/L, cT1 804ms, and PDFF 7.8%. Across treatment groups, baseline characteristics were broadly similar. The main statistically significant between-group difference was MASLD prevalence, which was highest in the IL-17i group (80%) compared with TNFi (47%) and csDMARD (20%). At follow-up, MDA was achieved by 12 patients: 8/8 in the TNFi group, 2/4 in the csDMARD group and 2/5 in the IL-17i group. New MDA achievement from baseline occurred in 10 patients, most frequently in the TNFi group (7/8), followed by IL-17i (2/5).

**Table 1:** Baseline and follow-up characteristics by treatment group.

|  | **csDMARD**<br>**Baseline**<br>**(n = 15)** | **csDMARD**<br>**Follow-up**<br>**(n = 15)** | **TNFi**<br>**Baseline**<br>**(n = 15)** | **TNFi**<br>**Follow-up**<br>**(n = 15)** | **IL-17i**<br>**Baseline**<br>**(n = 5)** | **IL-17i**<br>**Follow-up**<br>**(n = 5)** | **Whole**<br>**cohort**<br>**(n = 35)** |
| --- | --- | --- | --- | --- | --- | --- | --- |
| **Age (y)**<br>**[mean**<br>**(SD)]** | 50.8<br>(15.3) | - | 51.5<br>(11.4) | - | 40.0<br>(9.9) | - | 49.5<br>(13.3) |
| **Sex (Male)**<br>**[n (%)]** | 8 (53.3%) | - | 10<br>(66.7%) | - | 4<br>(80.0%) | - | 22<br>(62.9%) |
| **BMI [mean**<br>**(SD)]** | 27.6 (2.4) | - | 29.6<br>(5.0) | - | 30.4<br>(4.1) | - | 28.9<br>(4.0) |
| **Lean (<25**<br>**kg/m<sup>2</sup>) [n**<br>**(%)]** | 3 (20.0%) | - | 2<br>(13.3%) | - | 0 (0.0%) | - | 5<br>(14.3%) |
| **Overweight**<br>**(>=25 to**<br>**<30 kg/m<sup>2</sup>)**<br>**[n (%)]** | 9 (60.0%) | - | 7<br>(46.7%) | - | 2<br>(40.0%) | - | 18<br>(51.4%) |
| **Obese**<br>**(>=30**<br>**kg/m<sup>2</sup>) [n**<br>**(%)]** | 3 (20.0%) | - | 6<br>(40.0%) | - | 3<br>(60.0%) | - | 12<br>(34.3%) |
| **PsO [n (%)]** | 7 (46.7%) | - | 3<br>(20.0%) | - | 0 (0.0%) | - | 10<br>(28.6%) |
| **PsA [n (%)]** | 8 (53.3%) | - | 12<br>(80.0%) | - | 5<br>(100.0%) | - | 25<br>(71.4%) |
| **DAPSA**<br>**[mean**<br>**(SD)]** | 32 (21) | 26 (17) | 32 (20) | 11 (10) | 17 (3) | 10 (7) | 29 (19) |
| **-PASDAS**<br>**[mean**<br>**(SD)]** | 2.1 (1.0) | 1.5 (0.8) | 2.6 (0.9) | 1.5<br>(1.4) | 1.7 (0.3) | 0.8<br>(0.5) | 2.2<br>(0.9) |
| **-PASI**<br>**[mean**<br>**(SD)]** | 4.7 (3.8) | 0.8 (1.1) | 5.2<br>(10.3) | 0.8<br>(0.8) | 0.9 (1.0) | 0.4<br>(0.7) | 4.4<br>(7.2) |
| **-MASLD [n**<br>**(%)]** | 3 (20%) | 3 (25%) | 7 (47%) | 5 (42%) | 4 (80%) | 1 (33%) | 14<br>(40%) |
| **-MASH**<br>**(cT1 >= 800**<br>**ms & PDFF**<br>**>= 5%) [n**<br>**(%)]** | 2 (13.3%) | 4 (26.7%) | 5<br>(33.3%) | 6<br>(40.0%) | 2<br>(40.0%) | 1<br>(20.0%) | 9<br>(25.7%) |
| **ELF [mean**<br>**(SD)]** | 8.5 (0.8) | 8.8 (0.9) | 9.0 (0.9) | 9.2<br>(0.9) | 8.8 (0.7) | 9.1<br>(1.2) | 8.7<br>(0.8) |
| **Bilirubin**<br>**(umol/L)**<br>**[mean**<br>**(SD)]** | 9.6 (6.9) | 10.7 (5.8) | 11.5<br>(4.6) | 12.4<br>(5.9) | 11.2<br>(7.3) | 11.0<br>(5.4) | 10.7<br>(6.0) |
| **Albumin**<br>**(g/L) [mean**<br>**(SD)]** | 40 (4) | 41 (3) | 42 (4) | 42 (4) | 43 (4) | 45 (3) | 41 (4) |
| **ALT (IU/L)**<br>**[mean**<br>**(SD)]** | 31 (33) | 30 (16) | 36 (22) | 39 (35) | 38 (16) | 34 (19) | 34 (26) |
| **ALP (IU/L)**<br>**[mean**<br>**(SD)]** | 72 (28) | 81 (28) | 81 (14) | 80 (16) | 83 (27) | 95 (27) | 78 (23) |
| **CRP (mg/L)**<br>**[mean**<br>**(SD)]** | 2.5 (2.1) | 7.1 (14.9) | 5.5 (5.6) | 2.7<br>(2.2) | 3.2 (2.4) | 2.0<br>(1.1) | 3.8<br>(4.1) |
| **cT1 (ms)**<br>**[mean**<br>**(SD)]** | 770 (89) | 792 (150) | 800<br>(147) | 801<br>(141) | 919<br>(279) | 875<br>(287) | 804<br>(154) |
| **-PDFF (%)**<br>**[mean**<br>**(SD)]** | 5.6 (6.0) | 6.4 (7.7) | 7.8 (7.7) | 8.5<br>(8.0) | 14.0<br>(12.3) | 12.5<br>(13.1) | 7.8<br>(8.0) |

### IL-17 blockade is associated with improvements in liver disease

Over the follow up period, participants treated with csDMARDs showed numerical increases in both cT1 and PDFF (from mean of 770ms [SD 89] to 791ms [SD 150] and 5.6% [SD 6] to 6.4% [SD 7.7] respectively), which was also seen to a lesser extent with anti-TNF treatment (from mean of 800ms [SD 147] to 801ms [SD 141] and 7.8% [SD 7.7] to 8.5% [SD 8] respectively) (Table 1, Figure 1A-D). However, IL-17i reduced cT1 and PDFF (from mean of 919ms [SD 279] to 875ms [SD 287] and 14.0% [SD 12.3] to 12.5% [SD 13.1] respectively). Using a Bayesian mixed effects model, we found that only IL-17i had a meaningfully high probability of directional change (pd) in PDFF (−1.43, pd 89%) and cT1 (−39.8 ms, pd 93%), however their 95% credible intervals both included zero.

**Figure 1.**
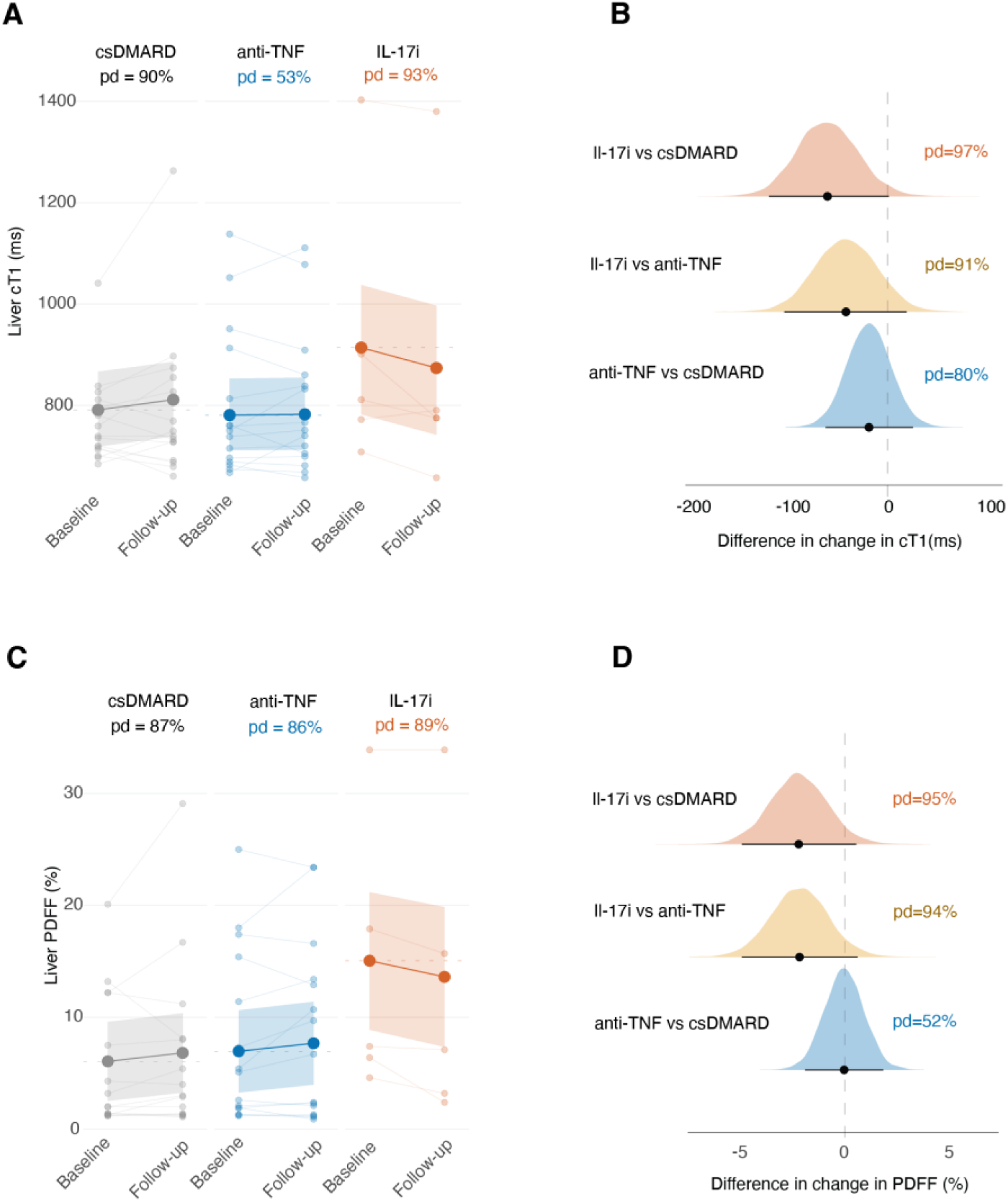
A. Liver MRI cT1 values (ms) at baseline, before initiation of new DMARD, and at follow-up, by treatment group. Lines represent individual patient trajectories. The band is the 95% CrI of the group mean. B. Posterior distribution of pairwise comparisons from A. 95% CrI is shown. C. Liver MRI PDFF values (%) at baseline, before initiation of new DMARD, and at follow-up, by treatment group. Lines represent individual patient trajectories. The band is the 95% CrI of the group mean. D. Posterior distribution of pairwise comparisons from C. 95% CrI is shown.

When considering the interaction between medication group and time, over the follow up period IL-17i reduced cT1 more than csDMARD by 60ms (CrI −121 to 3, pd 97%) and anti-TNF by 40ms (CrI −103.0 to 20.9, pd 91%). PDFF was reduced by 2.17% points more than csDMARD (CrI −4.85 to 0.56, pd 94%) and 2.16% points more than anti-TNF (CrI −4.90 to 0.59, pd 94%).

Exploring change in clinical features of PsD, both IL-17i and TNFi led to greater improvement in most measures compared to csDMARDs (Figure 2A). TNFi was associated with the largest reductions in DAPSA, SJC, and TJC. There was only minor correlation between clinical features and cT1 and PDFF when correcting for treatment group (Figure 2B), and the addition of clinical measures as covariates in the models for cT1 and PDFF did not improve them, suggesting that the effects we observed between treatment groups were not mediated by clinical state, but rather by the treatment itself.

**Figure 2.**
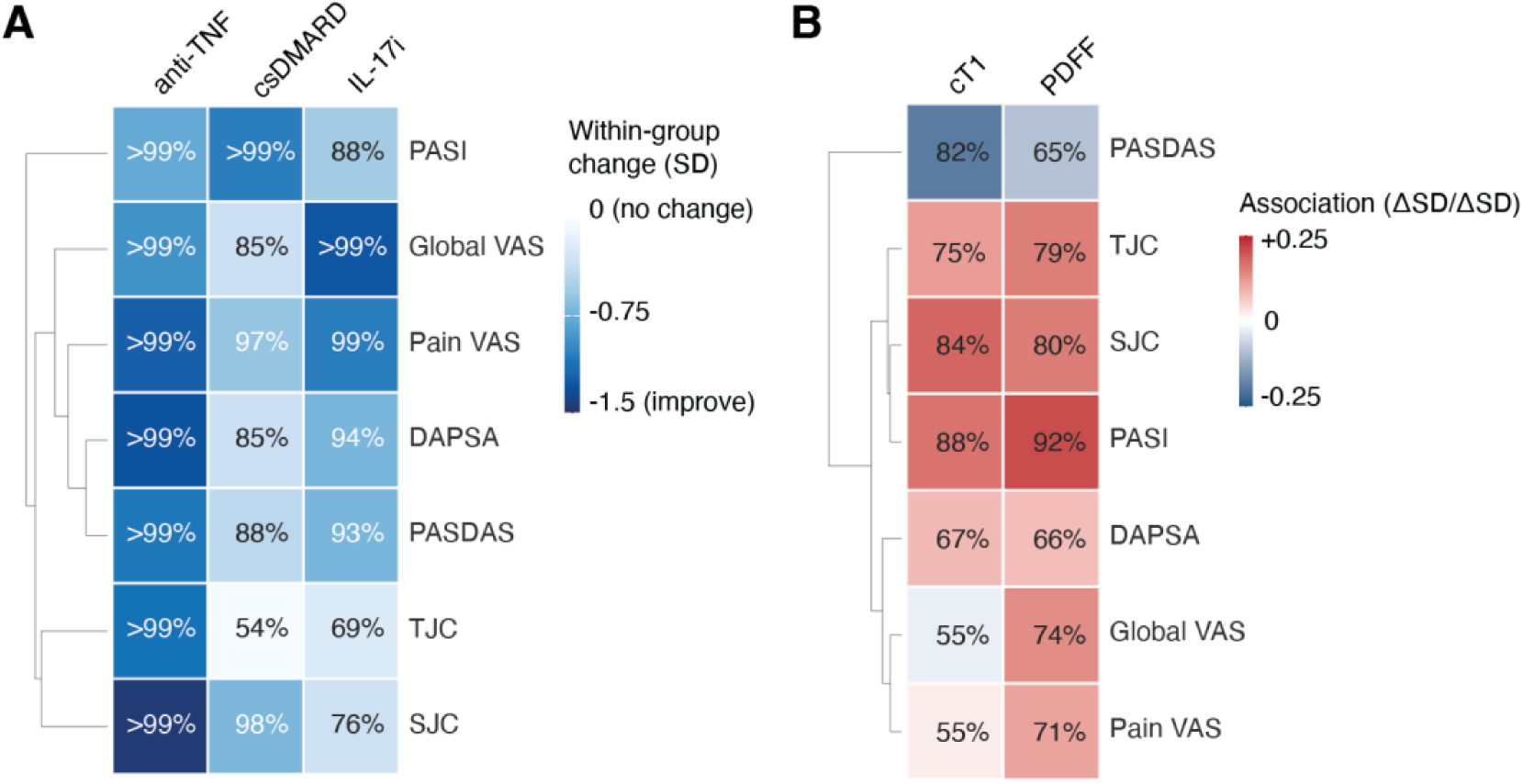
A. Heatmap showing within-group change from baseline to follow-up, for the indicated clinical measures. The scale represents effect size in standardised units (SD), and the number within the cell the probability of direction. B. Heatmap showing group-adjusted association between clinical measures and cT1/PDFF from baseline to follow-up. The scale represents association in standardised units (change/change), and the number within the cell the probability of direction.

### Liver disease covaries with proportions of circulating γδ and ThGM-CSF T cells

We next examined the association between cT1 and PDFF values at both timepoints, and T cell population frequencies in peripheral blood mononuclear cells (PBMCs), determined using flow cytometry, in a subset of 17 participants (4 on csDMARD, 8 on TNFi, and 5 on IL-17i). We detected 15 T cell subsets using surface markers (activated CD4^+^ T cells, naïve CD4^+^ T cells, CD4^+^ T effector, Th1, Th2, ThGM-CSF, Th9, Th17, Th22, T memory, regulatory T (Treg) cells, CD8^+^ T cells, MAIT cells, γδ T cells, and ɑβ T cells) with our panel (Supplementary Figure 2).[31] At baseline, and adjusting for treatment group, there was a strong positive association between ThGM-CSF proportion and liver PDFF values (pd 99% β=0.35 mean SD/SD, CrI [0.08, 0.64]) (Figure 3A). There was moderate evidence of a similar effect for cT1, although the credible interval included zero (pd 88% β=0.19 mean SD/SD, CrI [−0.13, 0.53]). We also observed high probability positive associations with γδ T cells and both cT1 and PDFF, and reciprocally, negative associations with ɑβ T cells.

**Figure 3.**
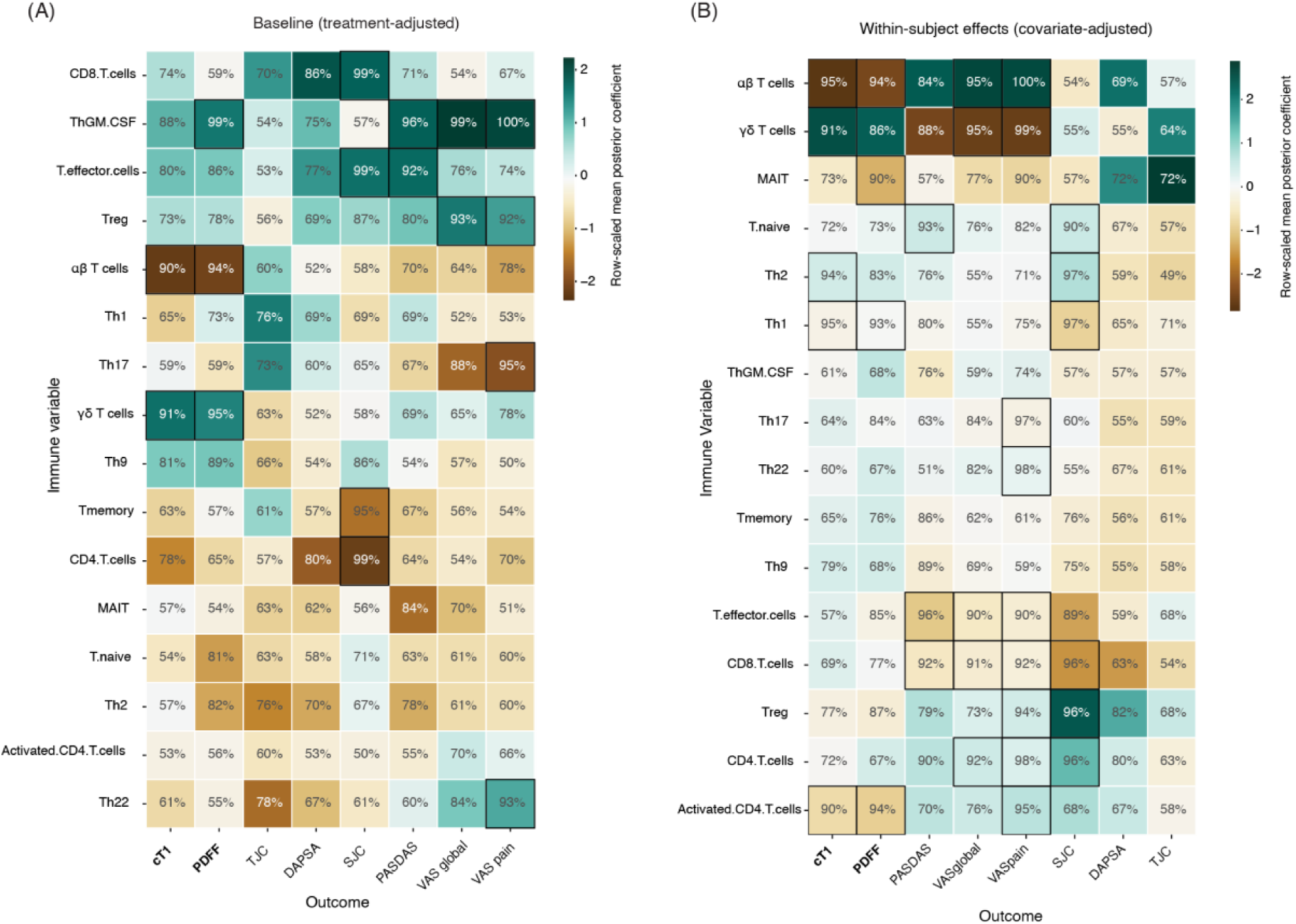
A. Heatmap of posterior coefficients for indicated associations (mean SD per SD), with row-scaled posterior probability of support indicated in text, at baseline. Black frames indicate posterior probability of directional difference ≥90%. B. Heatmap as in **A**, for within subject change over treatment period.

ThGM-CSF T cells were positively associated with VAS global and pain score, and effector CD4 and CD8 T cells positively associated with SJC. Treg cells were positively associated with VAS global and SJC.

We next explored how within-subject changes in T cell subset proportions over time predicted MRI and clinical parameter values at follow-up, adjusting for treatment group. We observed a positive association between increasing γδ T cell proportion and increasing cT1 and PDFF values at follow-up, and a reciprocal negative association with ɑβ T cells (Figure 3B). Increases in γδ T cells were associated with improvements PASDAS, and VAS global and pain scores at follow-up. Overall, flow cytometry demonstrated that γδ T cells were positively associated with increasing cT1 and PDFF over the follow up period, and at baseline ThGM-CSF T cells were positively associated with PDFF.

### Transcriptional differences in γδ and MAIT cells are associated with liver disease activity

We next performed single cell RNA sequencing of PBMCs in the same cohort of 17 patients, with pre- and post-treatment samples (Figure 4A). Given our flow cytometry data, we tested associations between T cell transcriptional profiles and liver MRI parameters (Figure 4B-E, Supplementary Figure 3). MAIT cells demonstrated the strongest transcriptional associations with cT1 at baseline. Higher cT1 was associated with increased expression of immediate early and cellular stress-response genes in MAIT cells, including *JUN* (standardised effect size (SES)= 16.21, FDR = 2.35 × 10^−13^), *DUSP1* (SES = 4.34, FDR = 2.41 × 10^−17^) and *PPP1R15A* (SES = 17.93, FDR = 9.44 × 10^−14^), together with genes consistent with cytotoxic/NK-like and tissue-positioning biology, including *CTSW* (SES = 10.23, FDR = 1.98 × 10^−14^), *KLRF1* (SES = 30.48, FDR = 2.38 × 10^−5^), *TCF7* (SES = 26.09, FDR = 1.03 × 10^−4^) and *GPR183* (SES = 37.62, FDR = 2.15 × 10^−3^). In contrast, higher baseline PDFF, indicating liver fat, was associated with lower expression of cytotoxic and immediate early-response genes in MAIT cells, including *GNLY* (SES = −3.81, FDR = 3.60 × 10^−30^), *FOS* (SES = −5.65, FDR = 1.08 × 10^−20^), *JUN* (SES = −2.82, FDR = 1.31 × 10^−13^), *DUSP1* (SES = −3.93, FDR = 4.00 × 10^−37^) and *PPP1R15A* (SES = −3.17, FDR = 4.24 × 10^−14^). These findings suggest that liver disease activity and steatosis are associated with divergent MAIT-cell transcriptional states at baseline, with higher cT1 linked to activation/stress and cytotoxic features, but higher PDFF linked to relative attenuation of these programmes.

**Figure 4.**
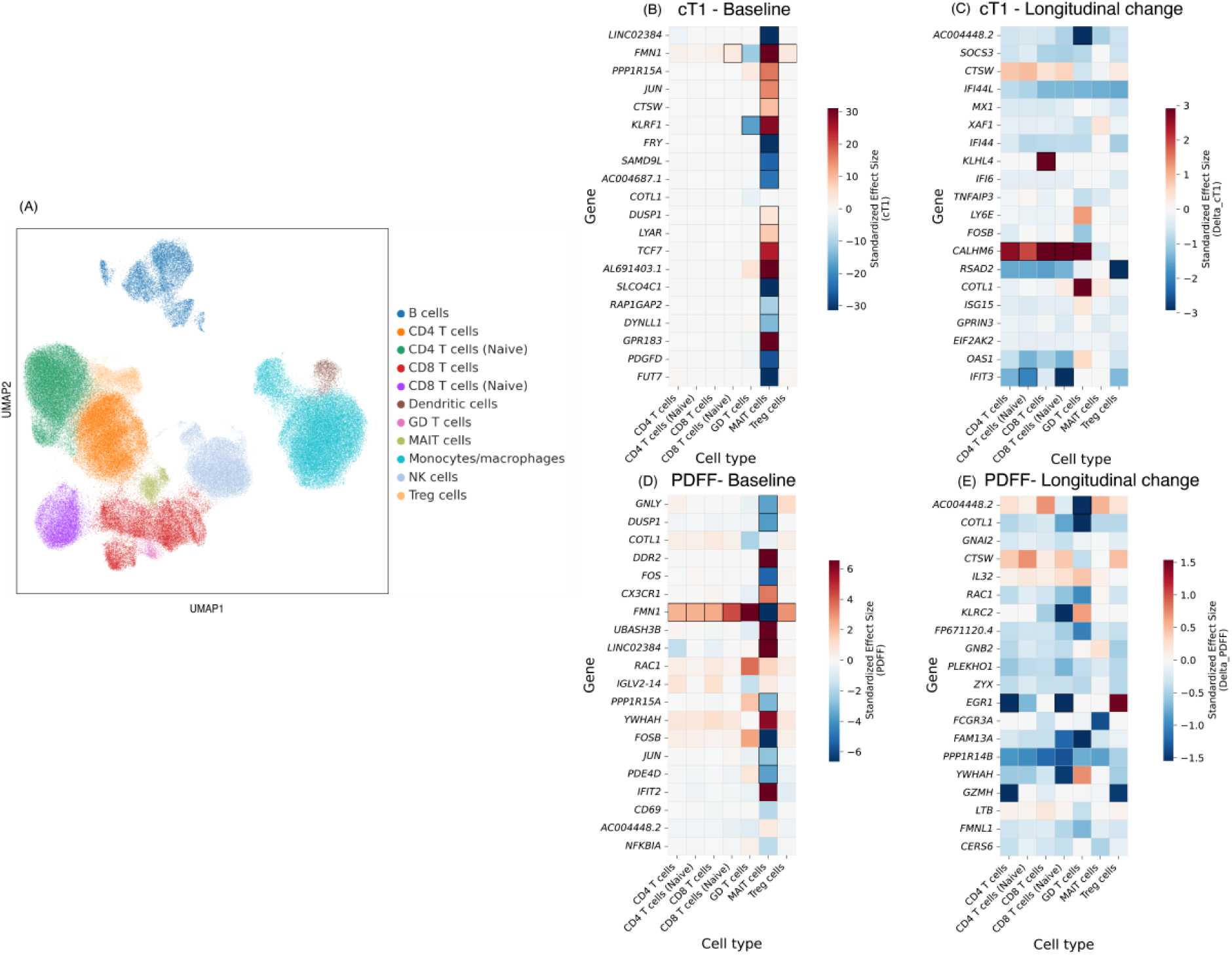
A. UMAP of 173,107 single cells integrated using totalVI (n=17 patients, with paired blood samples at baseline and follow up). B-E. Heatmaps show the top-ranked genes associated with cT1 and PDFF across T cell clusters at baseline (B and D), and showing paired longitudinal change (C and E). Differential expression was tested using Memento, modelling each MRI parameter as a continuous covariate and adjusting for age, sex, BMI and treatment group. Genes with FDR < 0.05 were ranked by a composite π-score incorporating statistical significance and effect-size magnitude, and the top 20 non-redundant genes were visualised. Colour denotes standardised Memento effect size. Black borders denote associations with FDR < 0.01 and absolute standardised effect size > 2.

Longitudinal analyses identified a set of baseline transcriptional features associated with subsequent change in liver MRI parameters. Increasing cT1 was associated with higher baseline *CALHM6* expression across CD4, naïve CD4, CD8, and naïve CD8 T cells, with the strongest signal in naïve CD8 T cells (SES = 5.14, FDR = 1.67 × 10^−5^). In contrast, decreasing cT1 was associated with higher baseline *IFIT3* expression in naïve CD4 T cells (SES = −2.09, FDR = 1.50 × 10^−15^) and naïve CD8 T cells (SES = −3.11, FDR = 8.12 × 10^−10^), consistent with an interferon-stimulated, activation-associated T cell state linked to subsequent improvement in liver disease activity. Change in PDFF showed fewer significant associations, but decreasing PDFF was associated with higher baseline *EGR1* expression in CD4 T cells (SES = −3.60, FDR = 2.63 × 10^−6^) and naïve CD8 T cells (SES = −6.35, FDR = 3.54 × 10^−8^).

## Discussion

In this exploratory prospective cohort study of 35 adults with PsD commencing new systemic therapy, only IL-17 inhibition was associated with reductions in hepatic steatosis (PDFF) and liver disease activity (cT1). Immune profiling from a subset of participants suggested that baseline γδ T cells and ThGM-CSF T cell abundance were associated with baseline PDFF and cT1 levels. We observed associations between within-patient changes in T cell populations and cT1 and PDFF values at follow up, adjusted for treatment group. Single cell RNA sequencing data also identified distinct transcriptomic signatures. Baseline liver disease activity was most closely linked to MAIT-cell activation, stress-response and cytotoxic/NK-like programmes, whereas steatosis was associated with relative suppression of these MAIT cell signatures. Following treatment change, γδ T-cell NK receptor genes showed opposing associations with cT1 and PDFF. Longitudinally, baseline *CALHM6* predicted increasing cT1, whereas interferon-stimulated *IFIT3*-associated naïve T-cell signatures and *EGR1*-associated signatures were linked to subsequent reductions in cT1 and PDFF, respectively.

cT1-associated upregulated genes included markers of T-cell activation, stress-response and inflammatory signalling, including *JUN*, *DUSP1*, and *PPP1R15A*, in keeping with an active immune-fibroinflammatory phenotype. *JUN* encodes c-Jun, a key component of the AP-1 transcription factor complex induced by T-cell stimulation, cytokine signalling, and MAPK pathway activation. *DUSP1* encodes a phosphatase that regulates MAPK signalling and may represent a counter-regulatory response to inflammatory activation; while PPP1R15A encodes *GADD34*, a regulator of the integrated stress response. Together, these genes suggest that cT1 may be associated not only with immune-cell abundance, but also with T cell activation, signalling, and cellular stress response.

In contrast, PDFF showed negative associations with cytotoxic and T cell activation-related genes, including *GNLY*, *FOS*, *JUN, DUSP1 and PPP1R15A*, suggesting that hepatic fat accumulation may lead to a more lipid-rich metabolic environment in which T-cell activation is comparatively reduced. *GNLY* encodes granulysin, a cytotoxic molecule expressed by T and NK cells. *FOS*, like *JUN*, forms part of early AP-1 transcriptional response to immune stimulation. These findings suggest that cT1 and PDFF capture overlapping but biologically distinct aspects of liver disease in PsD: inflammatory-immune activity and steatosis, respectively. Notably, the opposite direction of association observed for *JUN*, *DUSP1*, *PPP1R15A*, *FMN1*, and *LINC02384* with cT1 and PDFF in MAIT cells at baseline further supports this distinction.

Upregulated *GPR183* in MAIT cells at baseline was associated with higher baseline liver disease activity (cT1). *GPR183* acts as chemotactic receptor to guide migration of immune cells. Genome wide association studies have identified *GPR183* single nucleotide polymorphisms associated with susceptibility to inflammatory bowel disease (IBD).[32] Higher expression of GPR183 in IBD patients is associated with increased rates of skin psoriasis.[33]

Longitudinally, higher baseline *IFIT3* expression in naïve CD4 and naïve CD8 T cells was associated with subsequent reduction in cT1. *IFIT3* is an interferon-stimulated, activation-associated transcript that may reflect a T cell state shaped by innate cytokine signalling. Higher baseline *EGR1* expression was associated with subsequent reduction in PDFF. EGR1 is an immediate early transcription factor induced by cellular activation, stress and growth-factor signalling, and may reflect rapid transcriptional adaptation to inflammatory or metabolic cues. These findings suggest that baseline T cell transcriptional state may have relevance to subsequent improvement in liver inflammation and fat.

The improvements in liver disease MRI biomarkers seen in the IL-17 inhibitor subgroup is intriguing and biologically plausible. Increased Th17 cells and expression of IL-17A are markers of progression from MASLD to MASH.[34–36] Therefore, blockade of this axis could potentially exert direct or indirect beneficial effects on hepatic fat accumulation and fibro-inflammatory disease activity. Given the modest IL-17i sample size (n=5), these observations should be treated as hypothesis-generating but are consistent with previous studies showing improvement in liver health with IL-17i interventions. A Spanish prospective cohort study observed an improvement in liver stiffness, measured by transient elastography using Fibroscan, in 10 patients with psoriasis treated with secukinumab for 10 years.[37] A Japanese retrospective cohort study of 65 PsD patients treated with IL-17i reported significant decreases in FIB-4 and NAFLD fibrosis scores after 6 months of treatment.[38]

Several overall mechanistic insights emerge from the immune profiling that lend credence to the role of IL-17 in changes to liver disease features within the context of co-prevalent PsD. Mixed-effects modelling implicated higher ThGM-CSF and γδ T cell abundance with MRI biomarkers of liver disease. Our findings are in keeping with literature that demonstrate γδ T cells facilitate MASH and MASH-associated fibrosis.[39] This population of unconventional T cells produce cytokines that regulate inflammation, pathogen clearance, and tissue homeostasis.[40] A population of IL-17A secreting hepatic γδ cells have been identified that are sustained through the gut microbiota facilitating their activation, survival and proliferation.[41] Preclinical models suggest that γδ T cells may contribute to MASLD progression through IL-17A–dependent mechanisms. In diet-induced steatohepatitis, γδ T cells expand and adopt a more IL-17A-skewed phenotype, while loss of γδ T cells attenuates hepatic steatosis[42]. This highlights the key role of γδ T cells in liver microinflammation, through IL-17A mediated inflammation.

In keeping with our findings of associations for MAIT cell expression profiles and liver cT1, there is a recognised proinflammatory and profibrotic role of MAIT cells in the development of liver fibrosis in pre-clinical models. [20, 43, 44] Pharmacological and antibody-mediated inhibition of MAIT cells limit and regress fibrosis in MASH in human liver slices *in vitro* and mouse models.[45] MAIT cells have been shown to be dysfunctional in patients with MASLD, which is driven by accumulation of polyunsaturated fatty acids (PUFAs) in MAIT cells that leads to a state of metabolic exhaustion characterised by compromised mitochondrial respiration and glycolysis.[46] Raychaudhuri *et al.* have shown that MAIT cells are enriched in the synovial fluid of PsA patients compared to PBMCs, with a CD8⁺ predominance with upregulation of the IL-23 receptor.[47] This highlights the IL-17/IL-23 axis and MAIT cells in the pathogenesis of PsA. The therapeutic potential of MAIT cells is emerging, and they are being explored as candidates for novel immunotherapies such as chimeric antigen receptor (CAR)-MAIT cells.[48]

The study has several important strengths. We used paired, within-patient assessments combining validated MRI biomarkers of liver disease activity (cT1) and steatosis (PDFF) with detailed clinical phenotyping and multi-modal immunophenotyping (flow cytometry and single-cell RNA sequencing). This integrated design allowed us to examine clinical, imaging, and cellular correlates of treatment response in the same individuals — an approach that complements population-level observational data and offers mechanistic insights that cross-sectional studies cannot provide.

We must acknowledge several limitations of this study. The cohort size is modest, and treatment groups were small and non-randomised, limiting statistical power and raising the risk that observed differences reflect chance or selection bias. The immunology analyses were performed on a subset of patients (n=17), so those results require further validation in prospective studies. Furthermore, GM-CSF was assessed by the proxy of extracellular activation markers rather than intracellular cytokine staining. Finally, MRI metrics are sensitive, non-invasive biomarkers but are surrogate measures; histologic confirmation (liver biopsy) is the gold standard for steatohepatitis and fibrosis staging and was not available here.

Future work should prioritise larger, ideally randomised studies that stratify or enrich for baseline hepatic steatosis/disease activity to more robustly test drug-specific effects on liver outcomes. Such studies should capture serial measures of weight, alcohol use, metabolic medications, and insulin resistance. Longer follow-up is also needed to assess whether short-term imaging changes translate into altered risk of progressive fibrosis and liver-related clinical outcomes.

In summary, in this real-world, multimodal exploratory cohort we observed that IL-17i may reduce hepatic steatosis and liver disease activity in adults with psoriatic disease and MASLD. Baseline Th-GMCSF, MAIT, and γδ T cell immune populations with distinct expression profiles were associated with liver disease activity and hepatic steatosis, respectively. These findings support further focused investigation into how different immunomodulatory classes impact liver parenchymal biology and raise the possibility that treatment choice could be tailored to extra-articular comorbidity profiles in psoriatic disease.

## Supplementary material

### Supplementary methods

#### PBMC preparation

Peripheral blood mononuclear cells (PBMCs) were isolated from EDTA-anticoagulated whole blood using SepMate™ density gradient centrifugation. Blood was diluted 1:1 with Dulbecco’s PBS (dPBS), layered onto density gradient medium in SepMate tubes, and centrifuged at 1200 × g for 10 min at room temperature. The PBMC-enriched layer was collected, washed twice with dPBS, and cells were counted after resuspension, with filtration if clumping was observed. PBMCs were pelleted and cryopreserved in fetal bovine serum (FBS) containing 10% DMSO at a density-adjusted volume. PBMC vials were stored in liquid nitrogen.

#### Thawing and counting

PBMC vials were thawed in a 37°C water bath and resuspended in 1% fetal bovine serum (FBS)-phosphate-buffered saline (PBS). They were centrifuged, resuspended twice, and then cells were counted using a Luna 8-channel slide and the LUNA FX7 automated cell counter.

#### Flow cytometry

A complete list of antibodies is included in supplementary table 2. All antibodies used in this panel were validated for flow cytometry by the vendor or in the literature. Cells were labelled with Zombie UV viability dye (Biolegend) and Fc Block (BD Biosciences) for 30 minutes on ice in 96-well V-bottom plates (Sarsedt). After washing, surface staining was performed in Brilliant stain buffer (BD Biosciences, 50% total staining volume) and fluorescence-activated cell sorting (FACS) buffer (2.5g Bovine Serum Albumin and 2ml EDTA (0.5M) in 500ml dPBS) for 30 minutes. Cells were then fixed in freshly prepared 4% paraformaldehyde at room temperature for 15 minutes. The cells were washed in FACS buffer and acquired on a Cytek Aurora (5-laser) spectral flow cytometer. Pilot experiments were performed to determine the most suitable single-stained references (UltraComp eBeads™ or cells) for individual antibodies. Reference controls were processed similarly to fully stained samples on the same plate. Acquired samples were unmixed using SpectroFlo and analysed with FlowJo software.

Raw FCS files were compensated using experiment-specific compensation matrices and gated according to a predefined hierarchical gating strategy. [31, 49] (Supplementary Figure 1). Briefly, cells were sequentially gated to exclude debris and doublets, followed by live cell gating and identification of CD3⁺ T cells. Within the CD3⁺ compartment, prespecified subsets were defined based on canonical marker expression. Gating boundaries were applied consistently across all samples and population frequencies were exported for downstream analysis.

#### Single cell RNA sequencing

Counted cells were resuspended in Eppendorf tubes on ice. The lyophilized panel (TotalSeq™-C Human Essential Cocktail, V1.0 (Part: 750004666| Lot: B444666)) was equilibrated to room temperature and added to each sample. Hashing antibodies (TotalSeq™) were added to respective samples for labelling and incubated for 30 minutes at 4°C. Chilled PBS + 1% BSA was added to each sample, spun down and this cycle repeated a further two times. The final tagged cell suspensions were processed using the Chromium GEM-X single cell 5’ v3 gene expression with feature barcoding technology for cell surface protein kit, which includes post GEM-generation clean up, cDNA amplification and DNA quantification. Approximately 5,000 cells per sample were loaded for sequencing. Libraries were quality controlled and sequenced by Novogene (Cambridge, UK) using the NovaSeq X Plus.

Single-cell RNA and protein counts were pre-processed using Scanpy, selecting the top 4,000 highly variable genes. To integrate both modalities and correct for technical batch effects, we trained a totalVI joint probabilistic model conditioned on batch and sample ID covariates. The resulting totalVI latent space was used for UMAP dimensionality reduction and Leiden clustering. We employed a two-step annotation strategy: first isolating Gamma-Delta T cells via high-resolution clustering, then re-clustering the remaining cells to identify broad immune lineages and finally recombining all subsets into a complete annotated dataset.

To identify the most robust transcriptomic signatures associated with cT1 and PDFF across T cell subpopulations, differential expression results from *Memento* were filtered and ranked.[41] Analyses were performed independently for cT1 and PDFF at baseline and follow-up, and for longitudinal change in these parameters from baseline to follow-up (ΔcT1 and ΔPDFF). Within each T cell cluster, *Memento* was used to estimate expression moments and test associations with each continuous liver MRI parameter using bootstrap-based hypothesis testing with 1,000 iterations. Models were adjusted for age, sex, BMI and clinical group. The resulting coefficients represented the change in relative gene expression associated with each MRI parameter, with p-values adjusted for multiple testing using the Benjamini–Hochberg false discovery rate (FDR) procedure.

For visualisation, genes with FDR < 0.05 were ranked using a composite π-score incorporating both statistical significance and effect size, calculated as −log10(FDR + 1 × 10−300) × |β|. For each MRI parameter and analysis state, the top 20 non-redundant genes across T cell clusters were selected. Gene–cell-type associations with FDR < 0.01 and absolute standardised effect size > 2 were highlighted with a black border.

## Data Availability

Data availability statement: Summary data is included in the manuscript or uploaded as online supplemental information. Anonymised individual patient data can be shared upon request or as required by law and/or regulation and/or governance by and within the rules of UK Biobank access with qualified external researchers. Approval of such requests is at the discretion of the study sponsors and is dependent on the nature of the request, the merit of the research proposed, the availability of the data, and the intended use of the data.

## Acknowledgements

The authors thank the patients, the investigators and their teams who took part in this study. The authors acknowledge the funding bodies supported this research.

## Ethics and patient consent

Ethics committee approval was granted by London - Bromley Research Ethics Committee (REF: 20/LO/0616). All patients provided written informed consent.

---

**Supplementary Figure 1.**
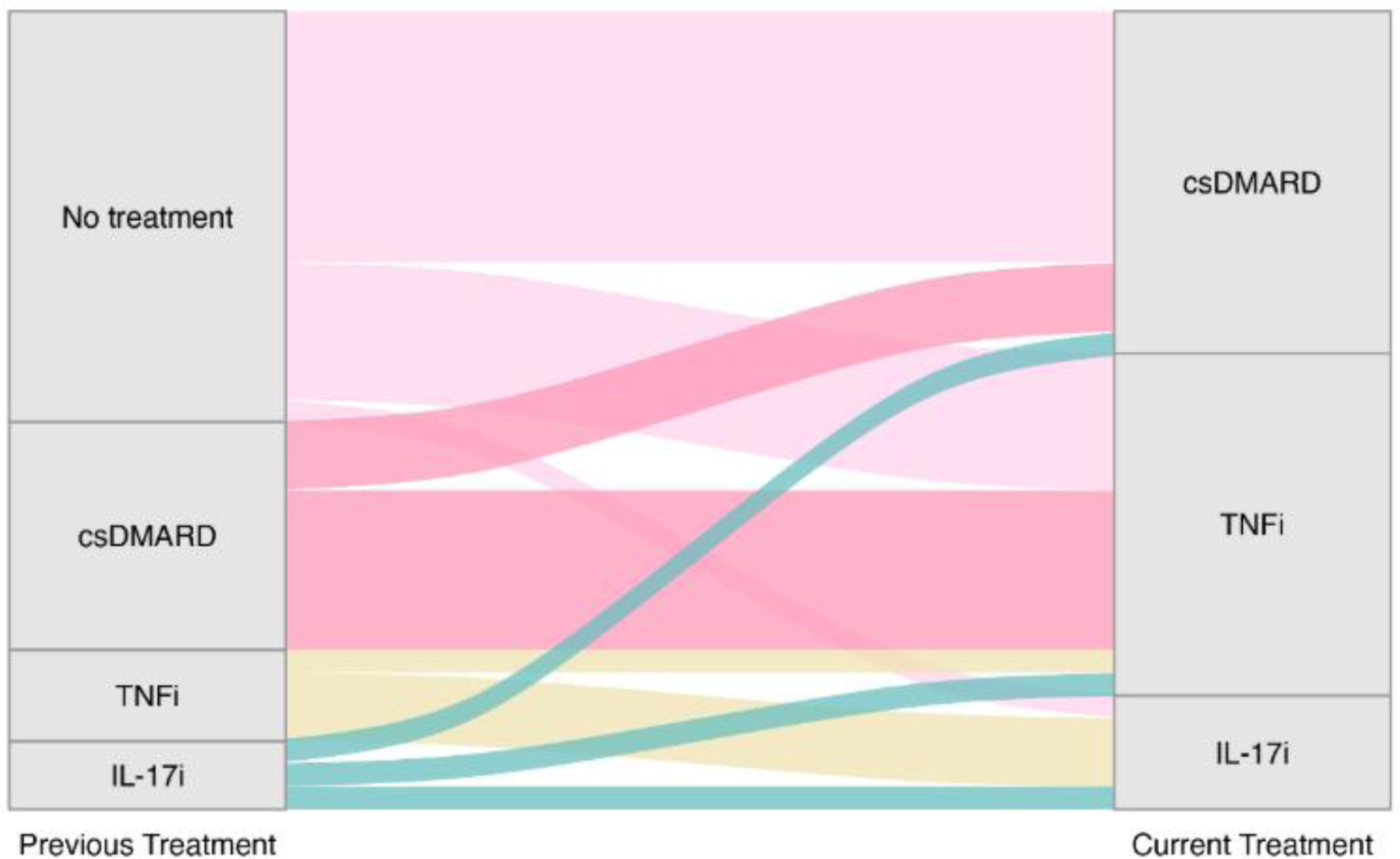
Sankey diagram to show treatment switches within the cohort. Previous treatment groups were ‘no treatment’ (n=18), ‘csDMARD’ (n=10), ‘TNFi’ (n=4) and ‘IL-17i’ (n=3). Follow-up treatment groups were ‘csDMARD’ (n=15), ‘TNFi’ (n=15) and ‘IL-17i’ (n=5). The majority (25/35, 71%) had PsA and a minority (10/35, 29%) PsO.

**Supplementary Figure 2.**
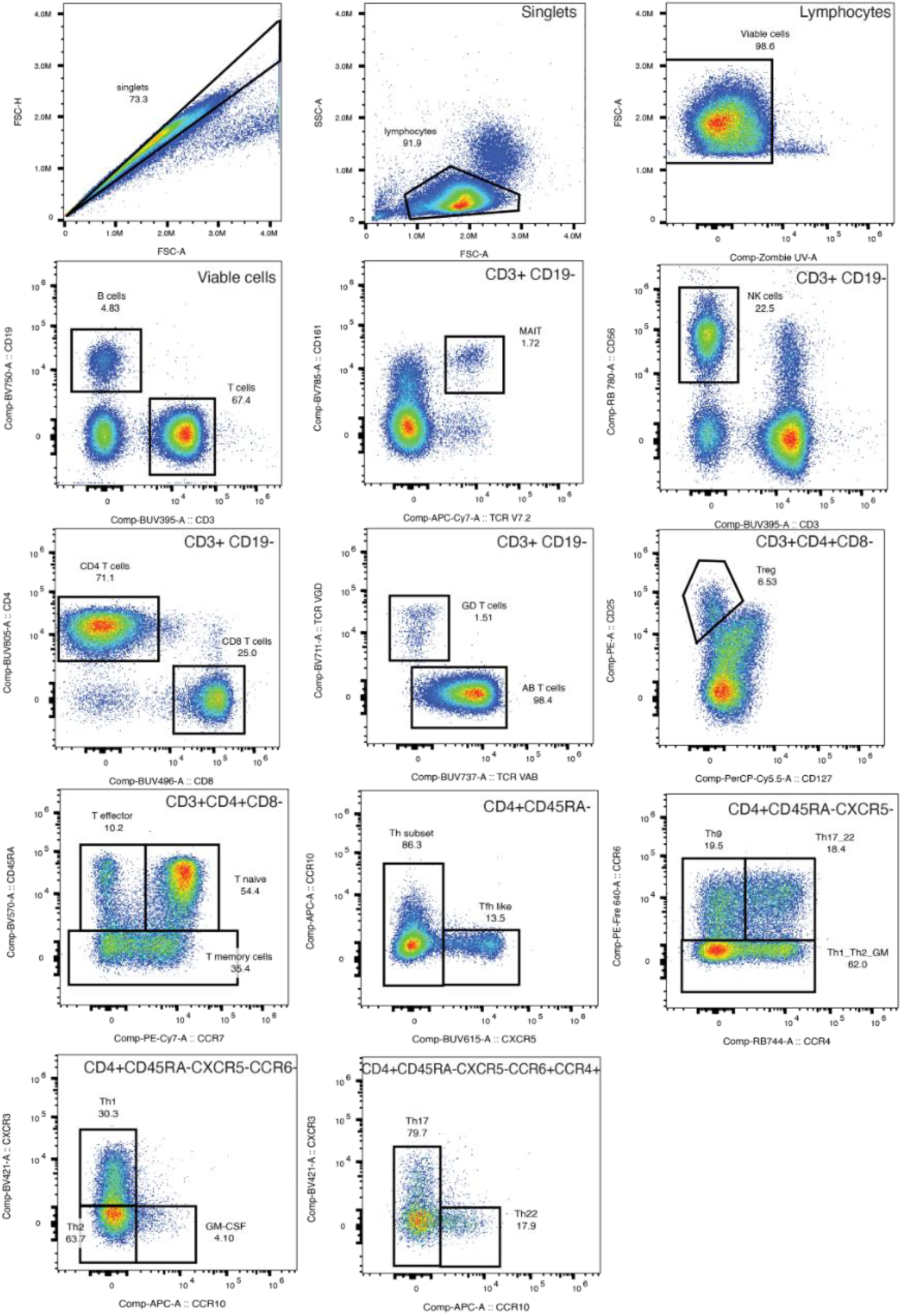
Gating strategy for flow cytometry analysis. Parent gating is shown, with indicated gates highlighting relevant populations.

**Supplementary Figure 3.**
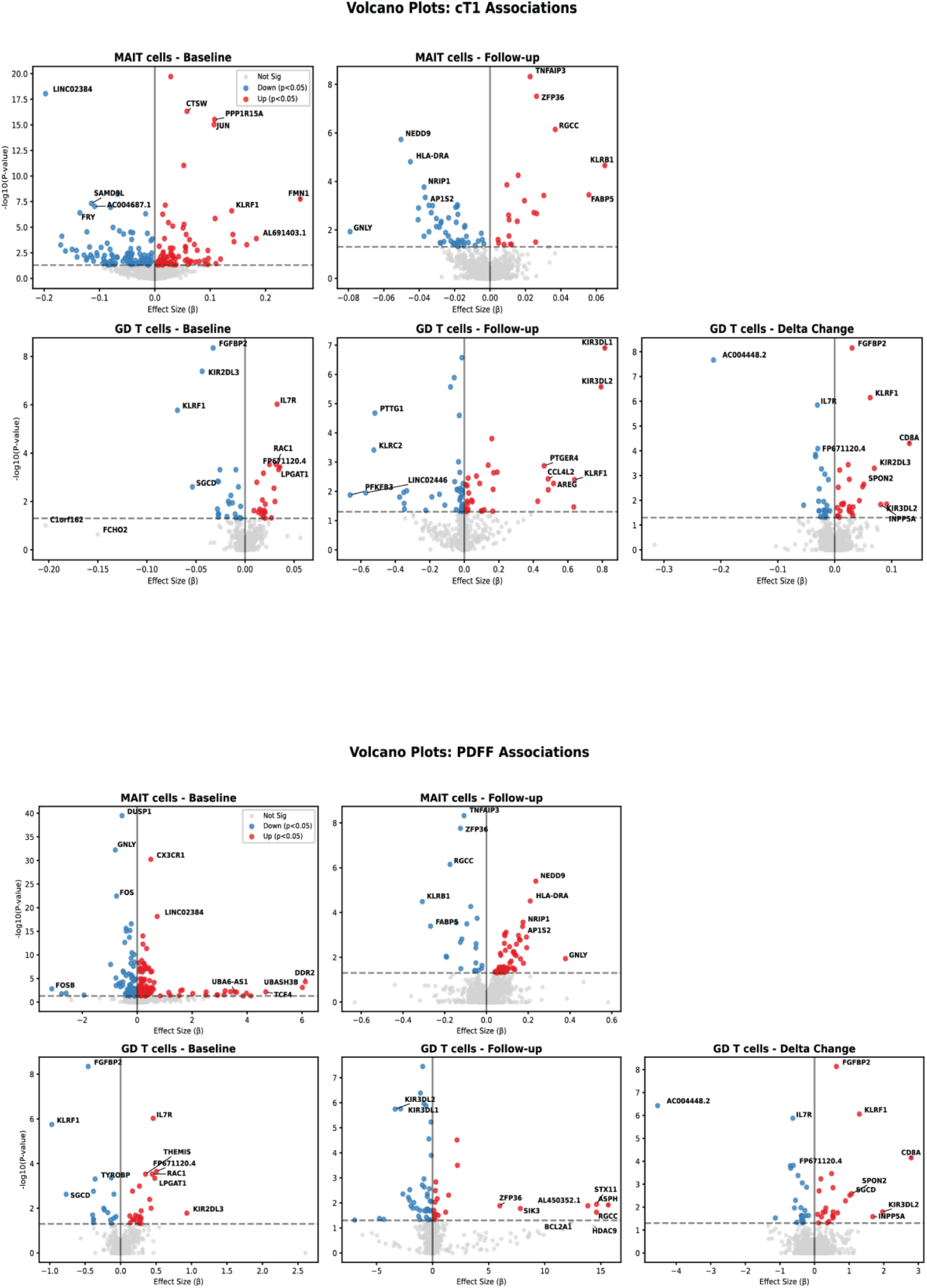
Volcano plots of top-ranked genes associated with cT1 and PDFF across cell clusters at baseline, follow-up and longitudinal change. Differential expression was tested using Memento, modelling each MRI parameter as a continuous covariate and adjusting for age, sex, BMI and treatment group.

**Supplementary table 1.** List of antibodies used in flow cytometry.

| Item Name |  |  | Catalogue # | Company |
| --- | --- | --- | --- | --- |
| Antigen | Fluor | Clone |  |  |
| Live-dead | Zombie UV |  | 423107 | Biolegend |
| CD4 | BUV805 | SK3 | 612888 | BD Biosciences |
| CD8 | BUV496 | RPA-T8 | 612943 | BD Biosciences |
| CD3 | BUV395 | UCHT1 | 563548 | BD Biosciences |
| TCR Vα $\beta$ | BUV737 | T10B9.1A-31 | 613014 | BD Biosciences |
| CD56 | RB780 | NCAM16.2 | 755586 | BD Biosciences |
| CD25 | PE | S20019C | 385607 | Biolegend |
| IgD | BV480 | IA6-2 | 566187 | BD Biosciences |
| CD38 | R718 | HB-7 | 567987 | BD Biosciences |
| CXCR5 | BUV615 | RF8B2 | 751293 | BD Biosciences |
| CD16 | BV650 | 3G8 | 302041 | Biolegend |
| CD27 | FITC | M-T271 | 356403 | Biolegend |
| CCR7<br>(CD197) | PE-Cy7 | G043H7 | 353225 | Biolegend |
| CD19 | BV750 | HIB19 | 302261 | Biolegend |
| CD14 | BUV661 | M5E2 | 741603 | BD Biosciences |
| CD11c | RB545 | Bly6 | 569252 | BD Biosciences |
| CD127 | PerCPcy5.5 | A019D5 | 351321 | Biolegend |
| HLA-DR | RB613 | G46-6 | 571123 | BD Biosciences |
| CD45RA | BV570 | HI100 | 304131 | Biolegend |
| TCR Vα7.2 | APC-Cy7 | 3C10 | 351713 | Biolegend |
| CD161 | BV785 | HP-3G10 | 339930 | Biolegend |
| TCR Vγ $\delta$ | BV711 | 11F2 | 568490 | BD Biosciences |
| CCR4<br>(CD194) | RB744 | 1G1 | 757578 | BD Biosciences |
| PD1 (CD279) | PE-dazzle594 | EH12.1 | 329939 | Biolegend |
| CCR6 | PEfire640 | G034E3 | 353448 | Biolegend |
| CXCR3 | BV421 | G025H7 | 353715 | Biolegend |
| CCR10 | APC | 6588-5 | 341505 | Biolegend |

